# Non-Congruent SARS-CoV-2 Waves in England

**DOI:** 10.1101/2021.01.25.21250440

**Authors:** Anthony J Brookes, Allyson Pollock, Danny Dorling

## Abstract

Examination of the chronology, location and size of waves of SARS-CoV-2 infection across England could shed light on the inter-play between the 1^st^ and the 2^nd^ Waves. From mid-October onwards such an analysis becomes increasingly difficult due to the emergence of a new strain (VOC-202012/01) and in light of the differential implementation of lockdown measures and tiers. Therefore, we sought to examine trends and correlations in virus prevalence and covid-related deaths spanning the start of the UK pandemic in March 2020 through to early November 2020 – i.e., including the first growth period of the 2^nd^ Wave. We found striking regional relationships between the 1^st^ and the 2^nd^ Wave that are difficult to explain other than by involving some role for changing levels of immunity in the population affecting the progression of the pandemic.

---

The first recorded death in England for which SARS-CoV-2 infection was listed as an underlying cause occurred on 30^th^ January 2020, in the South East of the country. The daily covid-19 death count began rising rapidly in March, leading to lockdown on the 23^rd^ of that month. The daily death rate peaked at 1279 on 8^th^ April 2020, by which time over 13,000 thousand people had died. Over 29% of these deaths occurred in Greater London^1^ despite only 16% of the population of England living in that region.

Mortality declined dramatically from April and throughout the summer to a low of 6 deaths per day on 3^rd^ September. This marked the end of England’s “1st Wave” of the pandemic. Thereafter, the “2nd Wave” began to emerge, growing more slowly than the first.

The regional distribution of infections in the 2^nd^ wave in England is markedly different from that in the 1^st^ Wave. Regions that suffered the very highest covid-19 mortality during the 1st wave have so far experienced smaller 2nd Waves. This was noticeable as early as mid-September, as we previously reported^2^, and our update of this analysis shows continuity of the pattern^3^.

A similar phenomenon has been noted elsewhere. For example, the largest US cities (e.g., New York) suffered a 1st Wave covid-19 death rate that rose quickly to high levels, whilst its 2nd Wave entailed far fewer deaths that peaked in August followed by a steady decline thereafter, as noted by the Financial Times Visual & Data Journalism team^4^. In contrast, small US towns and rural counties experienced a smaller 1st Wave mortality peak, followed by a 2nd Wave peak that was up to twice as high as the first and which persisted at those levels^4^. Similarly, municipalities in Italy’s Lombardy region that suffered the highest mortality during the 1st Wave are experiencing smaller 2nd Waves than other regions^5^. In this communication we extend our initial analysis of wave correlations in England^2^, using both infection and mortality data for the epidemic, examining wave size and rate of change.

## Community infection and death data

We used the weekly count of positive RT-PCR tests per Lower Tier Local Authority (LTLA) generated by Pillar 1 and Pillar 2 testing as a proxy for community infection. This testing has been scaled up massively since it began in early March, to around 340,000 tests per day by the first week of November (the time span of our analysis). To circumvent this distortion, we derived and employed an estimate of the percentage of tests that are positive per LTLA, and refer to this as the ‘Positive Test Rate’ (PTR). We also employed data on the percentage of deaths assigned to covid-19, as published weekly by ONS for each LTLA, and refer to this as the ‘covid-19 Death Rate’ (CDR). Using these measures we tested for correlations across the 1^st^ and 2^nd^ Waves over a range of different time windows. An online tool has been made available^3^ that presents more details of the data and the methods employed, and enable others to explore the robustness of our findings.

## Correlations between regions and between waves

Figure 1 shows how the PTR rose faster and higher in the 1^st^ Wave than in the 2^nd^ Wave. London was most severely affected during the 1^st^ Wave, followed by Northern regions. In contrast, during the 2^nd^ Wave the Northern regions were most badly affected, with London suffering relatively few infections. The same patterns are apparent in the CDR data^3^.

**Figure 1:**
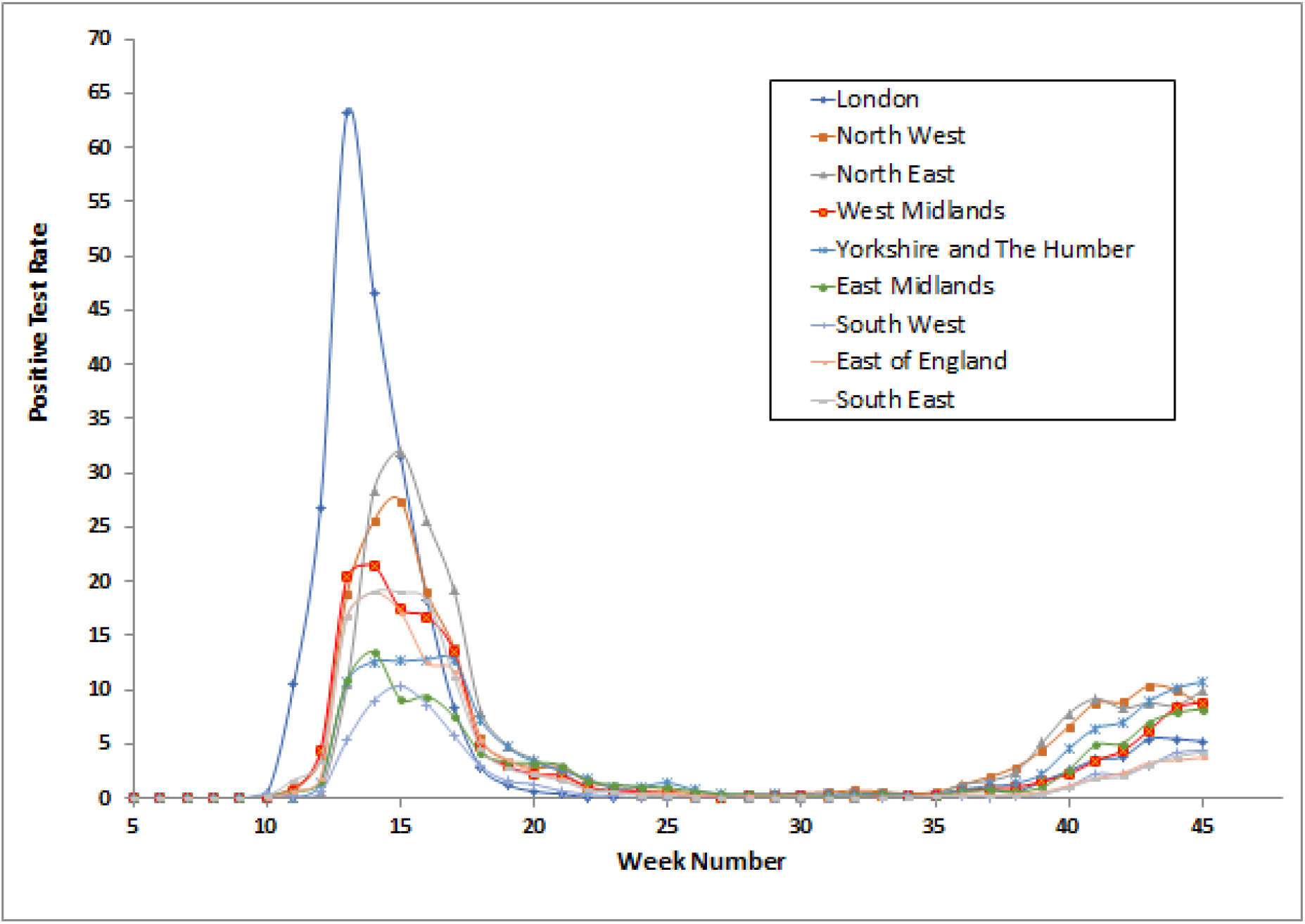
Combined Pillar 1 plus Pillar 2 Positive Test Rate (PTR) versus Week Number for England Regions.

## The scale of the 1st Wave affects the scale of the 2nd Wave

We plotted the 1^st^ Wave PTR against the 2^nd^ Wave PTR and CDR (Figure 2). This revealed: (a) regions with the smallest 1^st^ Waves experienced the smallest 2^nd^ Waves (e.g., South West LTLRs); (b) regions with progressively larger 1^st^ Waves experienced proportionally larger 2^nd^ Waves (e.g., Northern LTLRs); and (c) beyond a certain threshold of 1^st^ Wave size the 2^nd^ Wave size was markedly reduced (e.g., most London boroughs).

**Figure 2:**
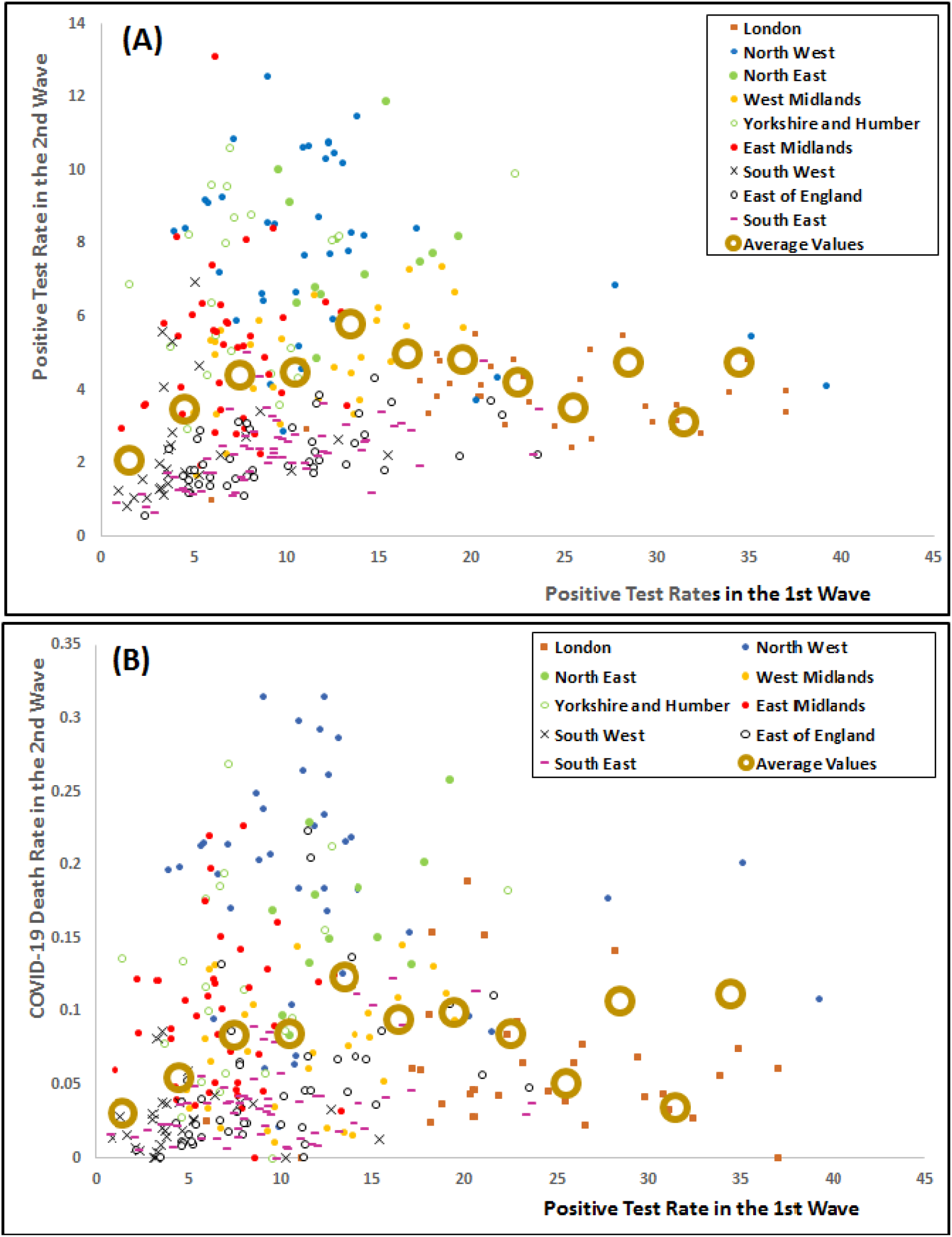
1st Wave Severity as a Predictor of 2nd Wave Severity. For each LTLA the average PTR (see Figure 1) was determined for the 1st Wave of the pandemic, spanning weeks 10-20 inclusive. For Panel (A) this was plotted against the average PTR per region for the 2^nd^ Wave over weeks 39-45 inclusive. For Panel (B) the positivity rate was plotted against the covid-19 Death Rate (CDR) per LTLA, i.e., the percentage of deaths recorded by ONS as being due to or involving covid-19, over 2^nd^ Wave weeks 39-45 inclusive. For each 3% window of PTR the 2^nd^ Wave data points were averaged (excluding isolated outliers) and plotted as large brown circles, to highlight the underlying trends in the data.

## The scale of the 1st Wave affects the rate of growth of the 2nd Wave

We plotted the 1^st^ Wave PTR against the rate of increase of PTR and CDR during the central growth period of the 2nd Wave (Figure 3). This revealed that regions of England with higher cumulative levels of past infection generally experienced slower rates of growth in both infections (PTR) and deaths (CDR) in the 2^nd^ Wave. A similar dampening of the rate of growth of PTR is apparent at the LTLA level. Unfortunately, this relationship could not be examined for CDR data at the LTLA level because the weekly death counts were too low and values below 3 are suppressed in the public datasets.

**Figure 3:**
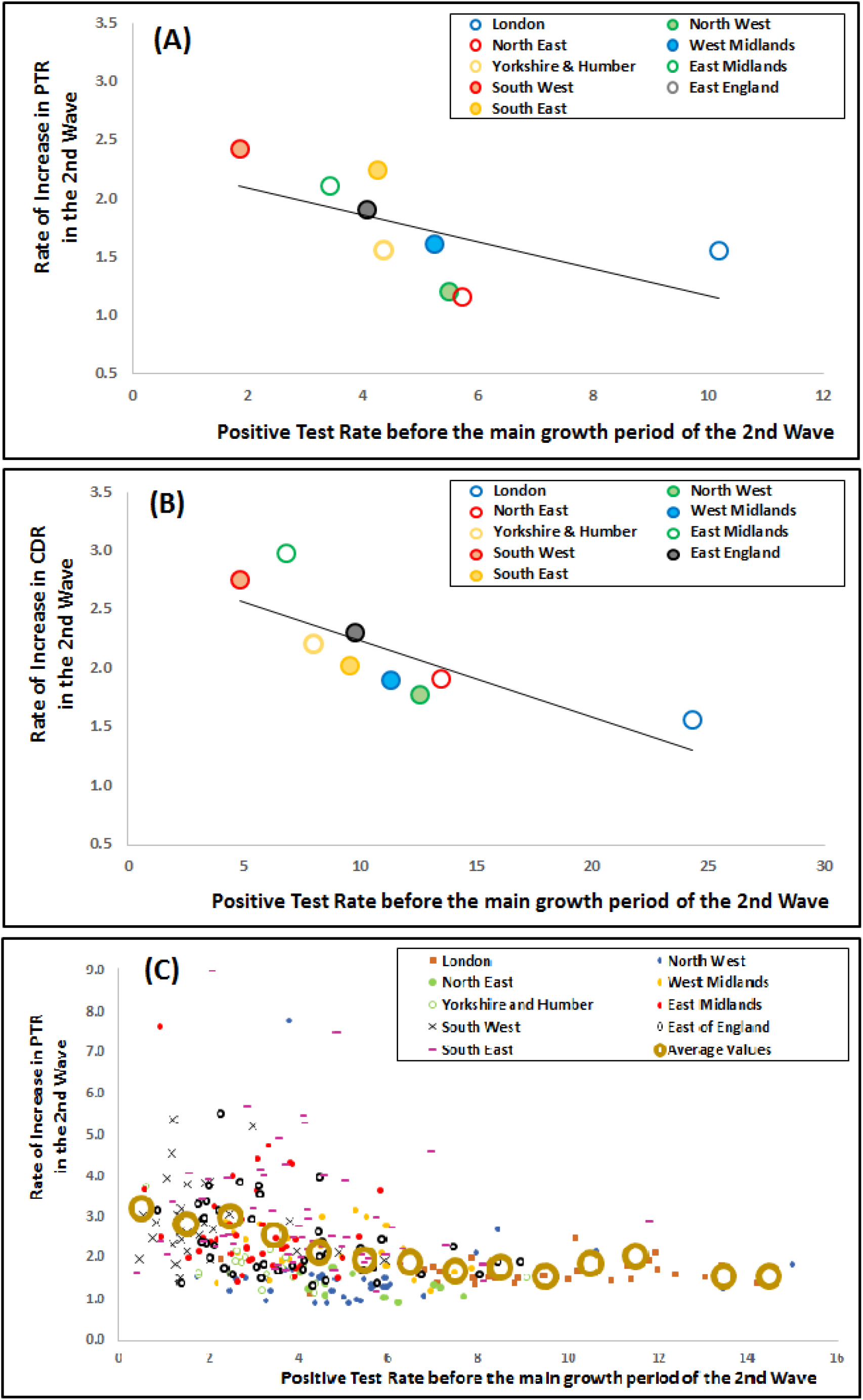
Accumulated Infections as a Predictor of 2nd Wave Growth Rates. Average PTRs (see Figure 1) were determined for large regions and LTLAs in England for weeks 10-38 inclusive, encompassing the 1^st^ Wave and the beginning of the 2^nd^ Wave of the pandemic. These were plotted against averaged 2-week increases occurring through weeks 39-45 inclusive, in: Panel (A), average PTRs for England regions, including a linear trend-line; Panel (B), average CDRs for England regions, including a linear trend-line; and Panel (C), average PTRs for LTLAs. For Panel (C), large brown circles indicate the average Y-axis values over each 1% window of X-axis values, and for clarity one outlier data point is not displayed (Brighton and Hove @ 2.76:17.56).

## Discussion

Our observations show that the size of the 1^st^ Wave of the SAR-CoV-2 pandemic in LTLA and broader regions of England predicts the size and rate of progression of the 2^nd^ Wave in those regions. Such correlations are also evident if one replaces the PTR data on the x-axis with CDR or ICU admission data, and all the signals are relatively insensitive to changes in the employed time windows. The online tool we have created^3^ enables people to explore these and other options.

When considering possible explanations for these observations, one might propose particular events such as the September and October opening of schools and universities, the imposition of Tiers in a few regions, and the onset of colder weather. However, these factors would arguably reduce rather than generate signals in the directions we observed. More extreme social distancing, shielding and other changes in behaviour consequential to experiencing a larger 1^st^ Wave could theoretically explain the noted patterns, as could changes (over time and place) in the demographics of the subpopulations sampled by testing. However, arguably, Track and Trace data do not support the notion that these are major factors.

An alternative explanation could involve dynamic changes in both individual and population levels of immunity – which both have a “continuum” rather than “all-or-nothing” character, as recognised in the classic introduction of the “herd immunity” concept^6^. Just as with other coronaviruses, SARS-Cov-2 infection is likely to lead to resistance to subsequent infection that is initially substantial (but never absolute) and which attenuates over many months ^7^,^8^,^9^,^10^,^11^, ^12^. At a population level the distribution of individual levels of immunity will provide some buffering of the spread of infection. That influence on viral transmission will be in the context of many other potentially relevant factors, such as seasonality, inward migration of non-immune cohorts, nutrition, hygiene, social-distancing, socio-economic factors, ethnicity, immunizations (including BCG), prior seasonal coronavirus infections, intervention implementations, and others. The relative impact of these myriad factors is hard to disentangle, but regardless, when a sufficient proportion of the population has had prior infection the resulting population immunity can be expected to lower - at least to some degree - the overall rate of viral transmission.

If population immunity was completely or largely absent from England (i.e., most or all of the population remains as vulnerable as in early 2020), the spring/summer lockdown would have suppressed virus prevalence to the same degree in all regions. Then, when lockdown was eased in late summer / autumn the pandemic would have resumed its growth from these scaled-down levels. Hence the expectation would have been that the virus would return in the 2^nd^ Wave with prevalence levels and rates of growth generally proportional to what was experienced in the 1st Wave. Likewise for the percentage of covid-19 related deaths. But this is not what occurred.

Instead, SARS-CoV-2 prevalence in regions that were most severely affected in the 1^st^ Wave fell to the very lowest of levels by the summer, and continued to have low levels relative to other regions as the 2^nd^ Wave unfolded. More generally, a greater 1^st^ wave severity led to a slower rate of growth in the 2^nd^ Wave. In Figure 2, a peak is apparent in the series of average values, such that LTLAs to the right of this (i.e., those with more infections in the 1^st^ Wave) do not suffer a more severe 2^nd^ Wave. This could be indicative of a level of prior infections whereby sufficient population immunity has been gained to buffer further transmissions of the virus. This would equate to most London boroughs, over half of all Northern regions, and a smaller fraction of other regions.

In conclusion, we have detected possible evidence of population immunity developing in many English regions, as of the end of week 45. Further analysis is warranted, in other contexts, with other datasets, and in more time periods. Whilst the level of immunity in the population could have been influencing the pandemic in some parts of England by early November 2020, other regions might not have been so well buffered. Subsequently, factors such as the onset of winter, the emergence of a new strain, changing severity of suppression measures, and the vaccination program, will be changing the amount of immunity needed to counter viral transmission across England.

## Data Availability

All data available via a website referred to in the manuscript

https://covidwaveexplorer.org/#intro

## Acknowledgements

The authors gratefully acknowledge valuable suggestions made by George Davey Smith (University of Bristol, UK) in the preparation of this manuscript, and have no conflicts of interest to declare.

## References

1. Deaths registered weekly in England and Wales, provisional - Office for National Statistics. https://www.ons.gov.uk/peoplepopulationandcommunity/birthsdeathsandmarriages/deaths/datasets/weeklyprovisionalfiguresondeathsregisteredinenglandandwales.

2. Why are coronavirus rates rising in some areas of England and not others? https://theconversation.com/why-are-coronavirus-rates-rising-in-some-areas-of-england-and-not-others-147160.

3. Brookes, A. J. covid-19 Pandemic Wave Analysis Tool. https://covidwaveexplorer.org/#intro

4. Covid-19: The global crisis — in data https://ig.ft.com/coronavirus-global-data/.

5. The valleys of the shadow of death - Italian towns hit hardest by covid-19 are doing better now | Graphic detail | The Economist. https://www.economist.com/graphic-detail/2020/10/31/italian-towns-hit-hardest-by-covid-19-are-doing-better-now.

6. Topley, W. W. C. Some Aspects of Herd Immunity. J. R. Soc. Promot. Health 56, 123–136 (1935).

7. Zuo, J. et al.. Robust SARS-CoV-2-specific T-cell immunity is maintained at 6 months following primary infection. BioRxoiv - Prepr. (2020).

8. Callow, K. A., Parry, H. F., Sergeant, M. & Tyrrell, D. A. J. The time course of the immune response to experimental coronavirus infection of man Nasal washings. 435–446 (1990).

9. W., N. K. et al. Preexisting and de novo humoral immunity to SARS-CoV-2 in humans. Science (80-.). 21, 1–9 (2020).

10. Grifoni, A. et al.. Targets of T Cell Responses to SARS-CoV-2 Coronavirus in Humans with covid-19 Disease and Unexposed Individuals. Cell 181, 1489-1501.e15 (2020).

11. Prior covid-19 infection offers protection from re-infection for at least six months | University of Oxford. https://www.ox.ac.uk/news/2020-11-20-prior-covid-19-infection-offers-protection-re-infection-least-six-months (2020).

12. Huang, A. T. et al.. A systematic review of antibody mediated immunity to coronaviruses: kinetics, correlates of protection, and association with severity. Nat. Commun. 11, (2020).

